# Pattern and Risk Factors for Nocturnal Enuresis among Children in Aseer Region, Southwestern Saudi Arabia: A cross-sectional study

**DOI:** 10.1101/2024.10.01.24314564

**Authors:** Youssef Ali Alqahtani, Ayed A. Shati, Ahmad A Alhanshani, Abdulaziz M. Al-Garni, Syed Esam Mahmood

## Abstract

Background

Nocturnal enuresis is defined as persistent bed-wetting at night beyond the age of five year old. To assess the prevalence and risk factors for nocturnal enuresis in children of the Aseer region.

**Materials and methods:** A descriptive cross-sectional survey via an online questionnaire was conducted targeting all accessible populations who are involved in childcare (5 to 18 years of age) in the Aseer region of Saudi Arabia. The questionnaire was circulated online using social media platforms by the research team. The questionnaire included items to address sociodemographic data of the child and the family, their medical history, and bed-wetting-related information.

**Results:** The study included 466 children, of which 145 (31.1%) complained of recurrent bedwetting. More than half of the children who experienced nocturnal enuresis were males (58.6%), and 31.7% were aged between 7-8 years, with a mean age of 5.9 years. A comparable proportion of fathers (59.3%) and mothers (57.9%) of the sampled children were university graduates. A total of 104 (71.7%) children started bed-wetting at 5 years of age, and 19.3% experienced bed-wetting from 6-7 years. Also, 126 (90%) children reported bed-wetting during sleep. Attention-deficit hyperactivity disorder, dark phobia, family troubles, and exposure to bullying were the most frequently reported risk factors.

**Conclusion:** Nearly 1 out of 3 children experienced bed-wetting, which is relatively common in boys than girls. Early toilet training and a supportive parental attitude toward bed-wetting is essential to improve the child’s quality of life.

## INTRODUCTION

Enuresis refers to the involuntary loss of urine during sleep that occurs at least twice a week in children older than 5 years of age (or the developmental equivalent) for at least 3 months, and it is the most common urologic complaint in pediatric patients. The International Children’s Continence Society has defined enuresis as wetting that occurs at night, whereas they no longer refer to daytime incontinence as diurnal enuresis.^1^ While the condition is not a serious health problem, it affects school-age children and even teenagers, and can be upsetting for the child as well as the family.^1,2^ Nocturnal enuresis is 2-3 times more common in boys than girls. About 20% of all children report occasional instances of bed-wetting until the age of 5 years, and up to 10% still have problems by 7 years of age. Also, an estimated 1-3% of teenagers report incidences of bed-wetting.^3–5^

Most children are fully toilet trained by 5 years of age; nevertheless, there is no specific date for developing complete bladder and bowel control. Often there are worrisome instances of bed-wetting for some children between 5 and 7 years of age, and a small proportion of children still wet the bed after 7 years of age.^6,7^ Therefore, parents should consult a pediatrician or any healthcare worker who is experienced in treating children with NE if the condition persists beyond 7 years of age, if the child starts to wet the bed after a few months of being dry at night, and if bed-wetting is accompanied by painful urination, unusual thirst, pink or red urine, hard stools, or snoring.^8^

Bed-wetting or enuresis is categorized as primary enuresis—urinary incontinence in a child who has never been dry, and secondary enuresis—incontinence in a child who has been dry for at least six months.^9^ Nocturnal enuresis in children is the second most reported pediatric problem after allergic diseases.^10^ Further, children with enuresis can develop a variety of behavioral, psychological, and social problems, including shame, blushing, lack of self-esteem, and aggression.^9–10^ Hence, it is important to identify the children at risk and implement therapeutic measures for treatment.

Many causative factors have been associated with bed-wetting, such as developmental differences, including the development of a child’s urinary sphincters, and diseases like diabetes and urinary tract infections. Emotional changes and conflicts, such as the birth of a new baby, scholastic or educational burden, and emotional crises, such as parental separation or divorce and family conflicts, are also reported as contributory factors.^11–12^ Having a global incidence of 1.4–28% among 6-12 years old children, the condition is troublesome for the families of these children, as it can result in considerable emotional distress and psychological consequences.^13–14^ In this light, the current study aims to assess the prevalence and pattern of nocturnal enuresis among the children of the general Aseer region population in southern Saudi Arabia.

## MATERIALS AND METHODS

A cross-sectional design was used to conduct this descriptive study through a survey. All accessible populations who are involved in pediatric care (for children below the age of 18 years) in the Aseer region of Saudi Arabia were invited to participate. Those participants above 18 years and those not willing to participate were excluded. Children with nocturnal enuresis secondary to other medical disorders or accompanying disorders like bowel and bladder dysfunction were also excluded. Clarifying the inclusion and exclusion criteria would help readers better understand the study population. The study was conducted in accordance with the Declaration of Helsinki. The Research Ethics Committee at King Khalid University provided ethical approval for the study (Approval number ECM#2020-0904), and data collection was started afterward with an electronic questionnaire. The questionnaire was circulated online by the researchers and their acquaintances through social media platforms seeking participation from eligible families with children to fill out the attached tool. The parents filled the questionnaires. Participation in this study was entirely voluntary, and confidentiality and anonymity were assured. Electronic informed consent was attached before the questionnaire in the provided links and the participants had approved before filling the questionnaire. The online questionnaire was designed by the research team under the guidance and consultation of a subject expert. The data collection was started on 1 July 2021 and continued till 31 December 2021. At the beginning of the questionnaire, a single question was put up enquiring whether the participating family had a child with recurrent bedwetting. In case of an affirmative response, the participant was directed to the subsequent questionnaire items, which covered the following items - socio-demographic data of the child and the family (age, gender, residential address, parents’ education, parents’ occupational details, and family history of chronic diseases); specific history of the child including chronic health problems, child-related stressors, environmental stressors, child abuse, and bullying; details of bed-wetting (age of onset, frequency, and effects on the child). According to Epi info Version, a sample size of 384 participants was estimated with a 5% margin of error and a 95% confidence interval ^15^. However, the statistical analysis and results were conducted on a sample of 466 children. A total of 500 people were invited. The response rate was 93.2%. Consistent reminders were employed to achieve this high response rate and address the possibility of non-response bias. To ensure the effectiveness and clarity of the questionnaire, it underwent a pilot test. Feedback, suggestions, and comments from the pilot test were used to refine and modify the questionnaire, making it more comprehensive and easily understandable for participants.

All statistical analyses were performed using SPSS version 22.0 (IBM Inc. Chicago, IL) using two-tailed tests, and a p-value < 0.05 was considered statistically significant. Descriptive analysis using frequency and percent distribution was done for all variables, which included the prevalence of bed-wetting among sampled families, demographic data of the children and their family, medical condition, family history of bed-wetting, bed-wetting data, risk factors, and consequences. Attention-deficit hyperactivity disorder (ADHD), dark phobia, family troubles, and exposure to bullying was diagnosed by a child and adolescent psychiatrist, and these children were under treatment for these conditions.

## RESULTS

Out of a total of 466 children, 145 (31.1%) children had instances of recurrent nocturnal enuresis. Out of these 145 children, more than half were males (58.6%; 85), and 31.7% were aged between 7-8 years, with a mean age of 5.9 ± 2.4 years old. Regarding the residential background, 96 (66.2%) children were from urban areas; further, 24.8% were the first-born child, while 37.9% were the fourth child or after. Perinatal complications during birth were reported for 19 (13.1%) children, and 15.9% had a chronic health problem, which was either constipation (6.2%), recurrent urinary tract infection (UTI) (3.4%), or sinusitis (2.8%) (Table 1).

**Table 1.** Demographic data of children with nocturnal enuresis in the Aseer region, Saudi Arabia.

| Demographic parameters | Frequency | Percentage |
| --- | --- | --- |
| **Gender** |  |  |
| *Male* | 85 | 58.6% |
| *Female* | 60 | 41.4% |
| **Age** |  |  |
| *5- 6 years* | 42 | 28.9% |
| *7-8 years* | 46 | 31.7% |
| *9-12 years* | 45 | 31.0% |
| *> 12 years* | 12 | 8.3% |
| **Residential background** |  |  |
| *Urban* | 96 | 66.2% |
| *Rural* | 49 | 33.8% |
| **Order of birth** |  |  |
| *1<sup>st</sup>* | 36 | 24.8% |
| *2<sup>nd</sup>* | 35 | 24.1% |
| *3<sup>rd</sup>* | 19 | 13.1% |
| *4th or more* | 55 | 37.9% |
| **Complications during delivery** |  |  |
| *Yes* | 19 | 13.1% |
| *No* | 126 | 86.9% |
| **Chronic health problems** |  |  |
| *None* | 122 | 84.1% |
| *Constipation* | 9 | 6.2% |
| *Recurrent Urinary Tract Infections* | 5 | 3.4% |
| *Sinusitis* | 4 | 2.8% |
| *Mental retardation* | 1 | .7% |
| *Urinary congenital anomalies* | 2 | 1.4% |
| *Epilepsy / neurological diseases* | 1 | .7% |

Table 2 presents the demographic data for families of the children with nocturnal enuresis – a comparable proportion of fathers and mothers (59.3% and 57.9%, respectively) of these children were university graduates. Ninety-seven children had working fathers (66.9%) children, while only 50 children (34.5%) children had working mothers; 12 children (8.3%) had lost their fathers. Regarding the family’s monthly income, 68 (46.9%) families earned between 5000 and 10000 SR monthly.

**Table 2.** Family characteristics of children with nocturnal enuresis in the Aseer region, Saudi Arabia.

| **Characteristic** | **Frequency** | **Percentage** |
| --- | --- | --- |
| **Father's education** |  |  |
| *Below secondary* | 24 | 16.6% |
| *Secondary* | 35 | 24.1% |
| *University/ more* | 86 | 59.3% |
| **Mother's education** |  |  |
| *Below secondary* | 30 | 20.7% |
| *Secondary* | 31 | 21.4% |
| *University/ more* | 84 | 57.9% |
| **Father's work status** |  |  |
| *Not working/ retired* | 36 | 24.8% |
| *Working* | 97 | 66.9% |
| *Deceased* | 12 | 8.3% |
| **Mother's work status** |  |  |
| *Not working/ retired* | 95 | 65.5% |
| *Working* | 50 | 34.5% |
| **Monthly income** |  |  |
| *< 5000 SR (1233 EUR)* | 38 | 26.2% |
| *5000-10000 SR (1233 EUR-2466 EUR)* | 68 | 46.9% |
| *10000-15000 SR (2466 EUR-3699 EUR)* | 39 | 26.9% |
| **Social status of parents** |  |  |
| *Married* | 124 | 85.5% |
| *Separated* | 14 | 9.7% |
| *Father deceased* | 7 | 4.8% |
| **Chronic health problem of parent** |  |  |
| *None* | 58 | 40.0% |
| *Diabetes Mellitus* | 66 | 45.5% |
| *Hypertension* | 33 | 22.8% |
| *Urinary Tract Infections* | 19 | 13.1% |
| *Renal problems* | 10 | 6.9% |

A total of 104 (71.7%) children started bed-wetting at the age of 5-6 years (Table 3) while 19.3% started between the ages of 6 and 7 years. Furthermore, 126 (90%) children reported bed-wetting during sleep. As for the frequency of bed-wetting, it was once every night for 62 (42.8%) children, 2-3 times per week in 43 (29.7%) children, and once a week in 17 (11.7%) children. Ninety-one (71.1%) children used the toilet before sleep, and 72 (49.7%) children had a habit of running to the toilet.

**Table 3.** Data related to bed-wetting in the 145 children.

| **Parameter** | **Frequency** | **Percentage** |
| --- | --- | --- |
| **Age of start bed-wetting** |  |  |
| *5 years* | *104* | *71.7%* |
| *6-7 years* | *28* | *19.3%* |
| *8-10 years* | *13* | *9.0%* |
| **Time of bed-wetting** |  |  |
| *All-day* | *14* | *10.0%* |
| *During sleep* | *126* | *90.0%* |
| **Frequency of bed-wetting** |  |  |
| *Once every night* | *62* | *42.8%* |
| *2-3 times/ week* | *43* | *29.7%* |
| *Once / week* | *17* | *11.7%* |
| *2-3 / month* | *10* | *6.9%* |
| *Once / month or more* | *13* | *9.0%* |
| **Does the child use the toilet before sleep time?** |  |  |
| *Yes* | *91* | *71.1%* |
| *No* | *10* | *7.8%* |
| *Do not know* | *27* | *21.1%* |
| **Does the child have to run to the toilet?** |  |  |
| *Yes* | *72* | *49.7%* |
| *No* | *53* | *36.6%* |
| *Do not know* | *20* | *13.8%* |

On analyzing the risk factors for bed-wetting in these children (Table 4), the most common child stressors were Attention Deficit and Hyperactivity Disorder (ADHD) (17.2%) followed by nyctophobia (dark phobia) (15.9%), stress and anxiety (11.7%), and living away from the father (9%). Regarding environmental stressors, being bullied at home by siblings was the most frequent (17.2%), followed by constant parental conflicts (16.6%), being bullied at school (13.8%), and moving out of or changing the house (13.1%). A total of 78 (53.8%) children were reported to hold their urine until the last second, and 31 (21.4%) were physically abused. Family history of nocturnal enuresis was reported in 20 (13.8%) children. Regarding the source of drinking water, 48.3% of the children reportedly consumed bottled water, while 47.6% of children consumed Saline Water Desalination Company.

**Table 4.** Risk factors for bed-wetting in the children of the Aseer region, Saudi Arabia.

| **Risk factors** | **Frequency** | **Percentage** |
| --- | --- | --- |
| **Child stressors/comorbidities** |  |  |
| *None* | 80 | 55.2% |
| *ADHD* | 25 | 17.2% |
| *Nyctophobia (dark phobia).* | 23 | 15.9% |
| *Stress & anxiety* | 17 | 11.7% |
| *Living away from father* | 13 | 9.0% |
| *Lack of attention* | 7 | 4.8% |
| *Depression* | 3 | 2.1% |
| *Social phobia* | 5 | 3.4% |
| **Environmental stressors** |  |  |
| *None* | 82 | 56.6% |
| *Being bullied at school by others* | 20 | 13.8% |
| *Continuous problems between the parents.* | 24 | 16.6% |
| *Moving out and changing the house.* | 19 | 13.1% |
| *Being bullied at home by siblings.* | 25 | 17.2% |
| *Moving out and changing school.* | 5 | 3.4% |
| **Does your child hold his urine until the last second?** |  |  |
| *Yes* | 78 | 53.8% |
| *No* | 28 | 19.3% |
| *Do not know* | 39 | 26.9% |
| **Is the child physically abused?** |  |  |
| *Yes* | 31 | 21.4% |
| *No* | 96 | 66.2% |
| *Do not know* | 18 | 12.4% |
| **Does any of the family members suffer from nocturnal enuresis?** |  |  |
| *Yes* | 20 | 13.8% |
| *No* | 107 | 73.8% |
| *Do not know* | 18 | 12.4% |
| **Source of drinking water** |  |  |
| *Wells* | 6 | 4.1% |
| *Saline Water Desalination Company* | 69 | 47.6% |
| *Bottled water* | 70 | 48.3% |

Table 5 tabulates the consequences of bed-wetting among children. A total of 37 (25.5%) children relapsed after being pertinent. When considering the academic performance of these children, it was found that only 4 children performed poorly (2.8%), while the academic performance was excellent in 60 (41.4%) children; 40 (27.6%) children were below the school-going age.

**Table 5.** Consequences of bed-wetting among children in the Aseer region, Saudi Arabia.

| Consequences of bed-wetting | Frequency | Percentage |
| --- | --- | --- |
| **Did your child's problem (bed-wetting) get relapsed after it was normal?** |  |  |
| *Yes* | *37* | *25.5%* |
| *No* | *108* | *74.5%* |
| **Academic performance** |  |  |
| *Below school-going age* | *40* | *27.6%* |
| *Poor* | *4* | *2.8%* |
| *Good* | *41* | *28.3%* |
| *Excellent* | *60* | *41.4%* |

Table 6. Distribution of frequency of bedwetting by children. Exact of 75.7% of children aged above 6 years had frequent NE compared to 64.3% of others aged less than 6 years with statistical significance (P=.049). Also, frequent NE was detected among 78.9% of 2nd children or more versus 52.8% of first child (P=.002). A total of 76.8% of children non-working mother complained of frequent NE in comparison to 64% of others for working mothers (P=.048). Other factors including child gender, social status of family, age of first NE and all were insignificantly associated with NE frequency.

**Table 6.** Distribution of frequency of bedwetting by children.

| Bio-demographic data |  | Frequency of Nocturnal Enuresis |  |  |  | p-value |
| --- | --- | --- | --- | --- | --- | --- |
|  |  | Frequent (nearly daily) |  | Infrequent (weekly) |  |  |
|  |  | N | % | N | % |  |
| Child gender | Male | 60 | 70.6% | 25 | 29.4% | .558 |
|  | Female | 45 | 75.0% | 15 | 25.0% |  |
| Child age | < 6 years | 27 | 64.3% | 15 | 35.7% | .049* |
|  | > 6 years | 78 | 75.7% | 25 | 24.3% |  |
| Child order | 1st child | 19 | 52.8% | 17 | 47.2% | .002* |
|  | 2nd or more | 86 | 78.9% | 23 | 21.1% |  |
| Child chronic health problems | No | 90 | 73.8% | 32 | 26.2% | .400 |
|  | Yes | 15 | 65.2% | 8 | 34.8% |  |
| Social status of parents | Together | 88 | 71.0% | 36 | 29.0% | .602<sup>#</sup> |
|  | Separated | 11 | 78.6% | 3 | 21.4% |  |
|  | Father dead | 6 | 85.7% | 1 | 14.3% |  |
| Father education | Below secondary | 19 | 79.2% | 5 | 20.8% | .719 |
|  | Secondary | 25 | 71.4% | 10 | 28.6% |  |
|  | University/ more | 61 | 70.9% | 25 | 29.1% |  |
| Mother education | Below secondary | 20 | 66.7% | 10 | 33.3% | .452 |
|  | Secondary | 25 | 80.6% | 6 | 19.4% |  |
|  | University/ more | 60 | 71.4% | 24 | 28.6% |  |
| Mother work | Not working/ retired | 73 | 76.8% | 22 | 23.2% | .048* |
|  | Working | 32 | 64.0% | 18 | 36.0% |  |
| Age of start NE | < 5 years | 78 | 75.0% | 26 | 25.0% | .301 |
|  | 5-7 years | 17 | 60.7% | 11 | 39.3% |  |
|  | 8-10 years | 10 | 76.9% | 3 | 23.1% |  |
| Time of nocturnal enuresis | All day | 9 | 64.3% | 5 | 35.7% | .533<sup>#</sup> |
|  | During sleeping | 91 | 72.2% | 35 | 27.8% |  |

*P*: Pearson $X^2$ test #Exact probability test
\* $P < 0.05$ (significant)

Table 7. Frequency distribution of bedwetting by risk factors of Nocturnal Enuresis (NE). Frequent NE was detected among 80.5% of children with no environmental stressors compared to 61.9% of others with stressors with recorded statistical significance (P=.013). Additionally, All other risk factors including child stressors, physical abuse, family history of NE, and source of drinking water were insignificantly associated with NE frequency among study children.

**Table 7.** Frequency distribution of bedwetting by risk factors of Nocturnal Enuresis (NE)

| NE risk factors |  | Frequency of NE |  |  |  | P-value |
| --- | --- | --- | --- | --- | --- | --- |
|  |  | Frequent (nearly daily) |  | Infrequent (weekly) |  |  |
|  |  | No | % | No | % |  |
| Child stressors | None | 58 | 72.5% | 22 | 27.5% | .979 |
|  | Yes | 47 | 72.3% | 18 | 27.7% |  |
| Environmental stressors | None | 66 | 80.5% | 16 | 19.5% | .013* |
|  | Yes | 39 | 61.9% | 24 | 38.1% |  |
| Does your child hold his urine until the last second? | Yes | 58 | 74.4% | 20 | 25.6% | .642 |
|  | No | 21 | 75.0% | 7 | 25.0% |  |
|  | Don't know | 26 | 66.7% | 13 | 33.3% |  |
| Is the child being physically abused? | Yes | 23 | 74.2% | 8 | 25.8% | .518 |
|  | No | 71 | 74.0% | 25 | 26.0% |  |
|  | Don't know | 11 | 61.1% | 7 | 38.9% |  |
| Does any of the family member suffer from a nocturnal enuresis? | Yes | 12 | 60.0% | 8 | 40.0% | .399 |
|  | No | 80 | 74.8% | 27 | 25.2% |  |
|  | Don't know | 13 | 72.2% | 5 | 27.8% |  |
| Source of drinking water for the family | Wells | 4 | 66.7% | 2 | 33.3% | .526<sup>#</sup> |
|  | Saline water conversion corporation | 53 | 76.8% | 16 | 23.2% |  |
|  | Bottled water | 48 | 68.6% | 22 | 31.4% |  |

*P*: Pearson $X^2$ test #Exact probability test
\* *P* < 0.05 (significant)

Table 8 represents the correctly prediction of bedwetting by children using significant risk factors of Nocturnal Enuresis (NE) by step wise binary logistic regression. Each step in the model represents the addition of a new variable to the previous step, indicating its influence on predicting bedwetting and the associated statistical significance.

**Table 8:**
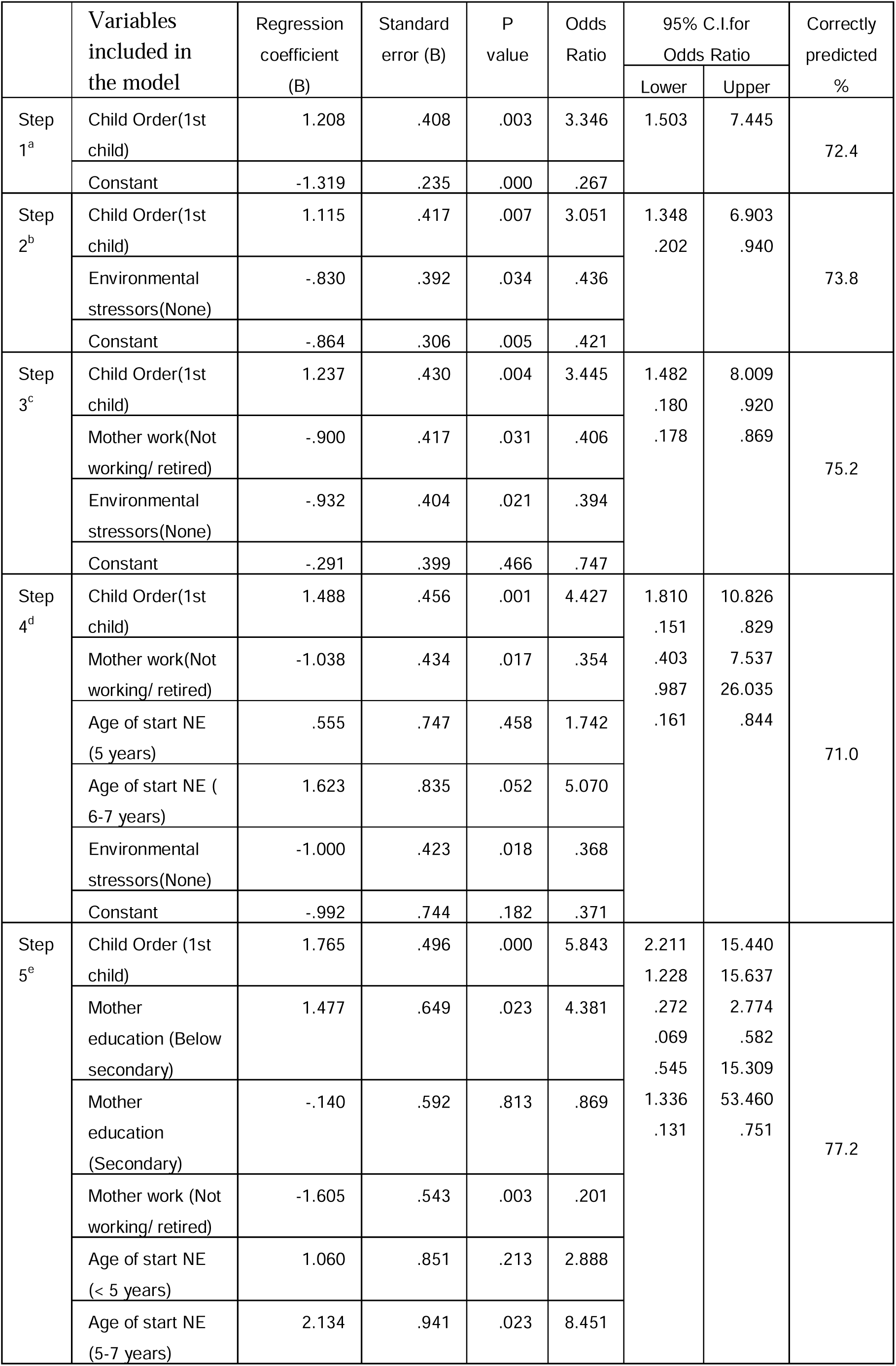
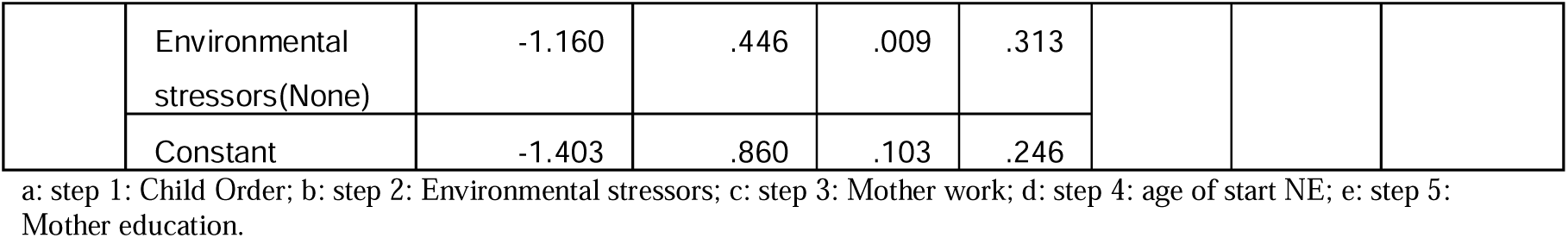
Forward stepwise likelihood model with correctly predicted of bedwetting by risk factors of Nocturnal Enuresis (NE)

The first step includes only one variable, “Child Order (1st child),” which has a p-value of 0.003. This indicates that there is a statistically significant association between being the first child and bedwetting. The odds ratio (O.R.) of 3.346 suggests that being the first child increases the likelihood of bedwetting by approximately 3.346 times. The model correctly predicted bedwetting in 72.4% of cases at this step.

In the second step, “Environmental stressors (None)” is added as an additional variable. Its p-value is 0.034, indicating a statistically significant association with bedwetting. The odds ratio is 0.436, implying that the absence of environmental stressors decreases the likelihood of bedwetting by approximately 0.436 times. The model now correctly predicts bedwetting in 73.8% of cases.

The third step introduces “Mother work (Not working/ retired)” as a new variable. It has a p-value of 0.031, suggesting a statistically significant association with bedwetting. The odds ratio is 0.406, indicating that not working or being retired reduces the likelihood of bedwetting by approximately 0.406 times. Additionally, “Environmental stressors (None)” remains in the model. The correct prediction rate increases to 75.2% at this step.

In this step, two variables related to the age of onset of bedwetting are added: “Age of start NE (5 years)” and “Age of start NE (6-7 years).” Both variables have p-values indicating statistical significance. The odds ratios suggest that the age of onset affects the likelihood of bedwetting differently depending on the range. The model also retains “Child Order (1st child)” and “Mother work (Not working/ retired)” from previous steps. The correct prediction rate decreases slightly to 71.0%.

The fifth and final step involves the inclusion of two variables related to “Mother education” and retains all variables from the previous steps. “Mother education (Below secondary)” and “Mother education (Secondary)” have p-values indicating statistical significance. The odds ratios show how different levels of maternal education affect the likelihood of bedwetting. The model now achieves a higher correct prediction rate of 77.2%.

Overall, this multivariate analysis table provides insights into the stepwise selection of variables and their associations with bedwetting. It suggests that factors such as child order, environmental stressors, mother’s work status, age of onset, and mother’s education play a role in predicting bedwetting.

## Discussion

The study revealed that one-third of children in our study sample complained of bed-wetting. Notably, there was significant variability in the estimated prevalence of NE among children in Saudi Arabia, ranging from 12% to 75% according to the study setting and the sampled age groups.^16–18^ The estimated prevalence of nocturnal enuresis in different regions of Iran is reported as 8.25% to 8.8%,^10–19^ compared to other studies conducted in Hong Kong, China, and Thailand^9,20,21^. Although the prevalence rate in Iran is higher, it is still lower than the reported prevalence in the Kingdom of Saudi Arabia, Turkey, and Burkina Faso.^22–24^ Furthermore, in our study, bed-wetting was more frequent among male children than females and in children aged 6-12 years old; this age group is older than that reported in the existing literature.^25,26^ Our finding is in-line with a previous survey in Saudi Arabia which found that the prevalence of NE were more in boys (62.9%) as compared to girls (37.1%).^27^ There is no specific reason for this gender difference. No significant difference among boys and girls was reported in an Egyptian study. ^28^ The current study also revealed that bed-wetting was more frequent among the first or last child. This phenomenon can be explained as an attention-seeking strategy adopted by older children who want to attract their parents’ attention after the birth of a younger sibling.

The current study also revealed that nearly three-quarters of the children wet their bed between the age of 5-6 years, as many children have incomplete toilet training by this age and the immature nervous system is unable to fully control the bladder.^29,30^ Regarding bed-wetting frequency, our results revealed that nearly half of the children (43%) did it every night, mainly during sleep time. This could be related to their urination behavior, as not all of them used the toilet before bedtime, and half of the children were habitual of running on feeling the urge to urinate, which means they waited until urgency (full bladder).

Regarding the risk factors analyzed among the current study in the children resorting to bed-wetting, child-related stressors were reported in almost half of the sampled children; it included ADHD, which is a major risk factor detected in different studies,^31–33^ dark phobia,^34,35^ stress and anxiety,^36,37^ family troubles and parental separation,^38^ exposure to bullying, and physical abuse.^39,40^ We also observed that few children had a positive family history of nocturnal enuresis, one of the main risk factors, besides the water source, which was bottled water in nearly half of the sample. Considering this relatively moderate proportion of the children who wet their bed, relapse rate after being dry was reported in only a quarter of these children. Notably, this did not affect the scholastic achievement of the school-going children, as only 3% had poor performance, while it was excellent in 42% of the children. In this study factors such as child order, environmental stressors, mother’s work status, age of onset, and mother’s education were found significant predictors to bedwetting. Similar significant associations between enuresis and child’s ages (years) P=0.002, residence P=0.002, and child order P=0.003 were found in another Saudi Study ^41^. Nocturnal enuresis was found to be associated with younger age, low socioeconomic and low educational level of the parents, non-working mothers, and family history of enuresis in an Egyptian study^28^. The following factors may have led to certain limitations in the present study. The cross-sectional nature of this study cannot confirm the causality association between the compared variables. The self-reported responses could over or underestimate the results. This was an online survey and not all children and parents have access to social media. In my opinion this is reflected by the fact that an important amount of parents had higher education levels. The authors did not use a voiding diary and evaluation of daytime symptoms in their study. We hope in future to have all the resources and less time constraints to do a comprehensive epidemiological survey and minimize the chances of the sampling bias. However the data included children of age groups 5–18 years and an extensive analysis has been made is the strength of our study.

## Conclusion

The study revealed that nearly 1 out of 3 children had experienced bed-wetting, which was relatively more among boys than girls. Attention-deficit hyperactivity disorder, dark phobia, familiar troubles, and exposure to bullying were the most frequently reported risk factors. Regardless of the estimated high prevalence rate, nocturnal enuresis had no effect on a child’s scholastic achievement. Overall, this study suggests that factors such as child order, environmental stressors, mother’s work status, age of onset, and mother’s education play a role in predicting bedwetting. The authors recommend early toilet training, a supportive parental attitude toward bed-wetting focusing on the child’s physical and mental health to improve the quality of life for children who experience bed-wetting.

## Data Availability

All data produced in the present study are available upon reasonable request to the authors

## Institutional Review Board Statement

The study was conducted in accordance with the Declaration of Helsinki, and approved by the Institutional Research Ethical Committee of the College of Medicine, King Khalid University for studies involving humans.

## Informed Consent Statement

Informed consent was obtained from all subjects involved in the study.

